# A multi-model analysis of risk factors influencing hippocampus degree centrality in Han Chinese adults who lost their only child

**DOI:** 10.1101/2024.11.22.24317808

**Authors:** Yang Cao, Min Guo, Yuxin Wang, Yifeng Luo

## Abstract

**Objective:** This study explores the relationship between Hippocampus Degree Centrality (DC) values and various risk factors in Han Chinese adults who have lost their only child, a phenomenon known as “shidu”. Our goal is to develop statistical models that have the potential to predict DC values and enhance clinical diagnosis and intervention strategies for Post-Traumatic Stress Disorder (PTSD) in these parents.

**Methods:** Participants were recruited from a PTSD survey conducted in Jiangsu Province, China, between September 2016 and March 2017. The survey included Han Chinese parents who had lost their only child to causes such as traffic accidents, explosions, suicide, cancer, and sudden death. Of the 237 participants, we focused on PTSD cases that met specific criteria, excluding individuals with adrenal disorders to eliminate confounding effects. After obtaining informed consent for follow-up assessments, we analyzed data from 109 eligible participants, examining a range of statistical models - linear, flexible, LASSO, ridge, and logistic regression - to identify correlations between DC values and various risk factors.

**Results:** Cortisol, gender, BMI, and memory capability are key contributors associated with DC values in Han Chinese adults who have experienced the trauma of losing their only child. We found that cortisol levels are negatively correlated with DC values in males, and are positively correlated in females (p-value = 0.017), which forms a gender-specific correlation. Additionally, BMI (p-value = 0.011) and memory scores (p-value = 0.157) indicate that physiological and cognitive factors can influence the hippocampus.

**Conclusion:** Linear and logistic models incorporating cortisol, gender, BMI, and memory capability show the greatest potential for clinical application in diagnosing DC abnormalities. These abnormalities are key indicators of stress levels and neurocognitive function in “shidu” parents, offering valuable insights for PTSD diagnosis and intervention strategies.

## Introduction

The one-child policy, which was in effect in China between 1979 and 2015, was a population planning policy implemented by the Chinese government to control the accelerating population growth in the country.^1^ It restricted Han Chinese families, the majority of the Chinese population, to having only one child. This policy, while addressing population control issues, simultaneously resulted in psychological and social challenges, particularly for Han Chinese parents who lost their only child.^2^ Known as “shidu” parents, these individuals were more likely to encounter trauma and stress related to the loss of their child. Such trauma and stress might make them vulnerable to some unhealthy psychological conditions, including the development of Post-Traumatic Stress Disorder (PTSD).

PTSD is a mental disorder that occurs and persists after experiencing serious threatening and catastrophic events. It is known to impair the brain’s physiology and functions, particularly within the hippocampus - a neuroanatomical region key to the stress response and cognitive processes like memory.^3^ Therefore, it is necessary to track the changes in the hippocampus in Han Chinese parents who lost their only child over time through their Hippocampus Degree Centrality (DC), a graph-based measurement of the brain network as well as a robust biological indicator of stress level and neurocognitive function.

Several other studies have explored the influence of various covariates on the hippocampus as well. For instance, in a study conducted by Mondelli et al.^4^, the researchers investigated the association between high cortisol levels and smaller left hippocampal volume. They utilized MRI to obtain the hippocampal volume and analyzed its relation with the cortisol level. While Mondelli et al. primarily addressed the physiological aspects of the cortisol-hippocampus relationship, our approach goes beyond by integrating advanced model selection techniques. This allows us to not only uncover nuanced cortisol-hippocampus dynamics but also has broader implications for clinical diagnosis and psychiatric interventions for “shidu” parents. Most importantly, although the above studies have explored the association between cortisol levels and hippocampal volume, especially in the context of psychiatric conditions such as schizophrenia, very few of them indeed studied the “shidu” parents, a very small but representative group with psychiatric concerns in China.

Building on these previous findings, our research compares several statistical models, including linear, flexible, LASSO, ridge, and logistic regression, to identify key factors that influence DC values in this unique population. Through this multi-model analysis, we aim to enhance clinical understanding and intervention strategies for PTSD in “shidu” parents, providing new insights into the role of the hippocampus in trauma-related disorders.

## Methods

### Participants

Participants for this study were recruited from a PTSD survey conducted in Jiangsu Province, China, between September 2016 and March 2017. The survey focused on Han Chinese parents who had lost their only child due to various causes, such as traffic accidents, explosions, suicide, cancer, and sudden death. The study was approved by the Medical Research Ethics Committee of Jiangsu University.^56^ In total, 237 parents participated, and we filtered the data to focus on PTSD cases^7^ that met specific criteria, including a prolonged duration since the loss, while excluding individuals with adrenal disorders (e.g., Addison’s disease or adrenal tumors) to eliminate confounding factors. After obtaining informed consent for follow-up physical assessments, we compiled a final dataset of 109 eligible participants. These individuals were assessed for various risk factors, including cortisol levels, immediate memory scores, age, sex, education, body mass index, and DC values.

### Missing Data Imputation

We conducted data preprocessing, which involved checking for inconsistencies, outliers, and missing data. To check for missing values, we found a total of 8 rows with at least one missing value for our 109 patients. For the variables, age, BMI, education, and memory each has 2 missing values, with the percentage of missing values in all variables < 5%. Although it is impossible to assess if the missing values are missing at random (MAR) statistically, we can observe that most of the 8 observations with missing values have cortisol values lower than 10. Thus, we can assume the missing values are MAR. The percentage of missing values for the 4 covariates is 1.8% which is less than 5%, so we can apply multiple imputations on the data^8^. To solve the missing value problem, we used multiple imputations of m = 5, which uses the average of 5 regression models, to fill in the missing values.

During the feature engineering phase, we used the average of the left and right DC values as our primary outcome variable. Since the roles of the left and right hippocampi in the brain’s overall function may not be entirely consistent, adopting the average value helps to better reflect the hippocampus’ overall functionality while mitigating the impact of hemispheric asymmetry. This approach offers a more balanced representation of hippocampal activity, which is crucial for understanding the effects of PTSD.

Using the average value not only enhances the comprehensiveness of our analysis but also reduces the influence of random measurement errors in individual data points, leading to a clearer reflection of the overall trend. Furthermore, as additional data samples are collected, a model based on average values will be more adaptable, facilitating the integration of new data without the need to differentiate between left and right origins. This flexibility ensures that the model remains scalable over time, improving its generalizability and long-term applicability.

### Model Selection

After deciding on the average DC value as our primary variable, we proceeded with the development of various models, including linear, flexible, LASSO, ridge, and logistic models, to analyze the relationships between DC values and various risk factors.

#### Linear Regression

For our linear regression model, we assessed the variable importance score (VIP) to conduct variable selection. For the retained key covariates, we employed a flexible modeling approach by incorporating quadratic terms, spline terms, interaction terms, and factorization terms for these four covariates. We then calculated and compared their Akaike Information Criteria (AICs)^9^ and selected the model with the smallest AIC value to be our optimal model.

#### Penalized Regression

Using the features in the optimal model selected by the smallest AIC values in the linear regression model, we then fit a Ridge regression and a LASSO model using the same features.

The penalization of the two models is selected by minimizing the Sum of Squared Errors (SSE) using a grid search method^10^.

#### Logistic Regression

Our original outcome metric is a continuous variable (mean of left and right DC values). We need to transform our outcome into binary variables for logistic regression. Since there is no existing dichotomization method mentioned in clinical documents, we categorized the DC values by the average values of DC. For example, observations of the upper 50% DC values are assigned to “1” and the lower 50% are assigned to “0”. Then, we initialized our model construction with all covariates and used p-values of coefficients to filter important ones.

## Results

### Sample Demographics

After conducting multiple imputations on the missing values, a total of 109 Han Chinese adults who lost their only child were included in our study. The demographics of study participants are summarized in Table 1.

**Table [1]:**
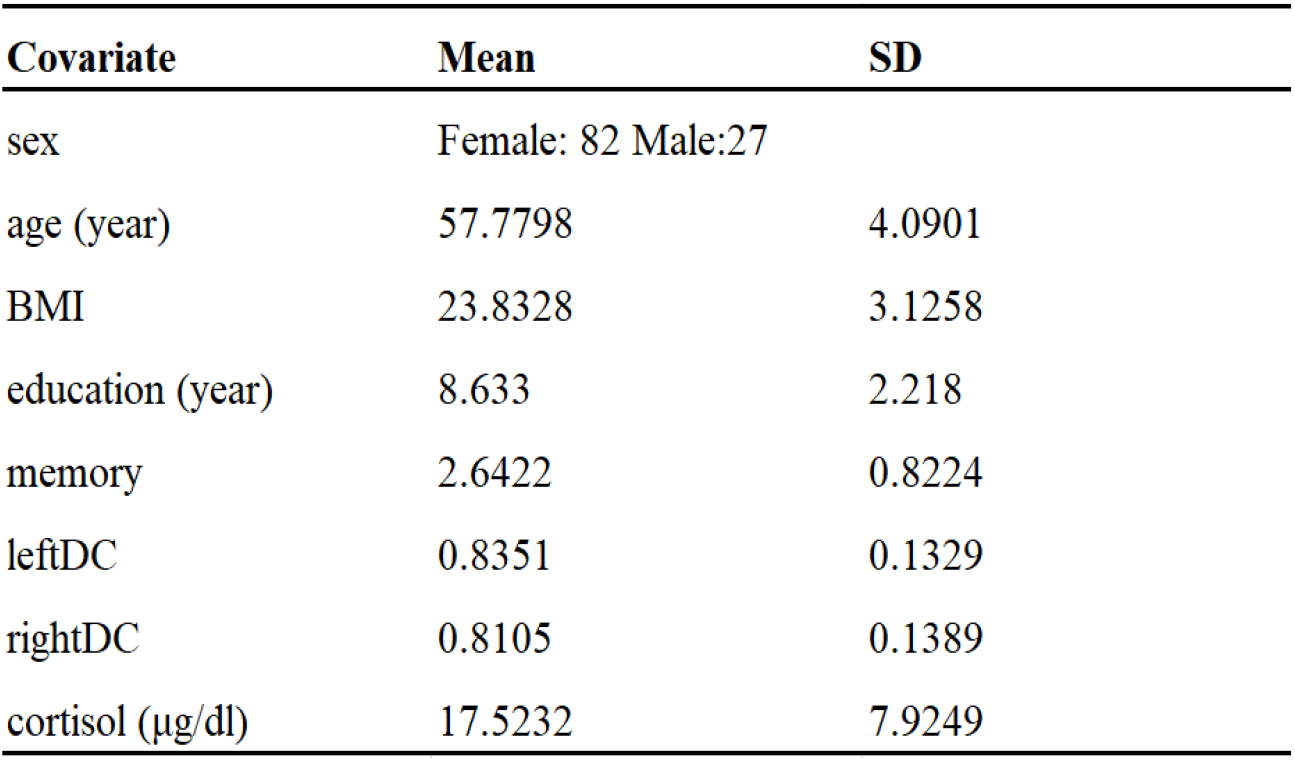
Demographics of study participants: Han Chinese adults who lost an only child.

| Covariate | Mean | SD |
| --- | --- | --- |
| sex | Female: 82 Male:27 |  |
| age (year) | 57.7798 | 4.0901 |
| BMI | 23.8328 | 3.1258 |
| education (year) | 8.633 | 2.218 |
| memory | 2.6422 | 0.8224 |
| leftDC | 0.8351 | 0.1329 |
| rightDC | 0.8105 | 0.1389 |
| cortisol (µg/dl) | 17.5232 | 7.9249 |

### Linear Regression

In our initial analysis, we assessed the VIP and excluded age and education level from further model exploration due to their exceptionally low VIP scores, as illustrated in Figure [1].

**Figure [1]:**
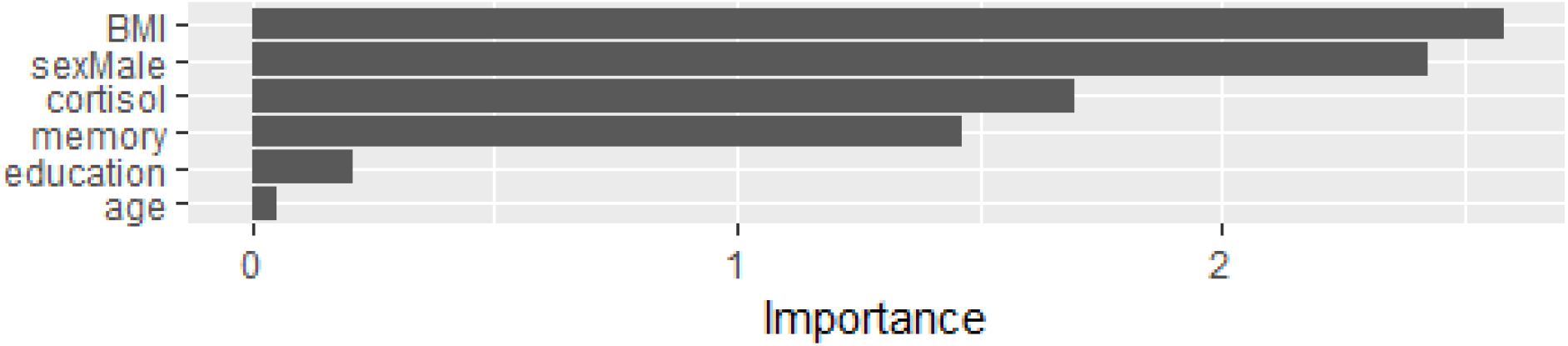
Variable importance plot (VIP) of all covariates.

Subsequently, we retained four key covariates - BMI, sex, cortisol, and memory score - for further modeling analysis. Employing a flexible modeling approach, we incorporated quadratic terms, spline terms, interaction terms, and factorization terms for these four covariates. All models are detailed in Figure [2], and their AICs are listed in Table[2]. In the model selection process, we compared the AIC between any successive models and chose the model with lower AIC. For instance, when comparing Mod1 and Mod2, we selected the reduced model (Mod2) without the interaction term between sex and BMI due to its lower AIC, then expanded the model to include other flexible terms, such as factorization of memory scores in Mod3. After evaluating eight models, we identified Mod2 as the optimal model due to its smallest AIC. Notably, we avoided a stepwise selection process, as it failed to account for the completeness of full terms for each variable. For example, in the comparison between Mod4 and Mod8, we preferred a result that either included or excluded all quadratic and cubic terms of cortisol, rather than selecting the cubic term while excluding the quadratic term, as often seen in stepwise procedures.

**Figure [2]:**
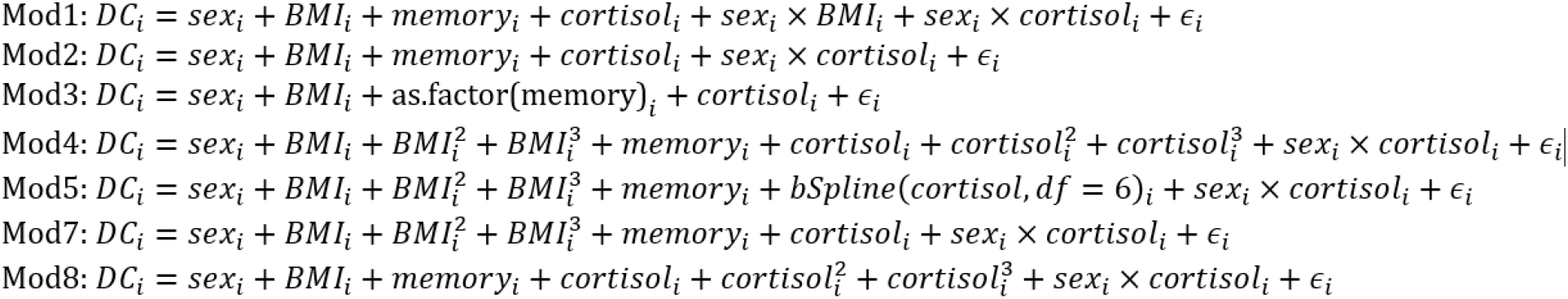
Model statement of 8 linear models.

**Table [2]:** AIC of 8 models.

| Model | AIC |
| --- | --- |
| mod1 | -156.8139 |
| mod2 | -158.7517 |
| mod3 | -157.3872 |
| mod4 | -156.6061 |
| mod5 | -153.3662 |
| mod6 | -153.3591 |
| mod7 | -158.391 |
| mod8 | -157.6298 |

Our findings provide a profound understanding of the relationship between DC values and various risk factors. We found that BMI (p-value = 0.011) and memory scores (p-value = 0.157) are negatively associated with DC values, while females’ cortisol levels (p-value = 0.017) are positively associated with DC values and males’ cortisol levels are negatively associated. These findings suggest that these risk factors may play a significant role in influencing the cognitive function of parents who have lost their only child. In detail, when holding all other covariates fixed, every 1 unit increase in BMI is associated with an average decrease of 0.0091 units in DC value. Similarly, every 1 unit increase in memory score is associated with an average decrease of 0.0189 units in DC values. For males, a 1 ug/dl increase in cortisol level is linked to an average decrease of 0.0034 units in DC values, while for females, it is associated with an average increase of 0.0036 units in DC values.

### Penalized Regression

After features were manually selected by scrutinizing the AIC as the model measurement, we employed two penalized methods to corroborate the initial feature selection. In Ridge Regression, an L2 penalty term of coefficients was introduced to the Sum of Squared Errors (SSE), and a grid search of lambda values was conducted to minimize the SSE. As shown in Figure [3], the Ridge Regression results demonstrated a gradual shrinkage of regression coefficients towards zero as the logarithm of lambda increased. Lambda was chosen as the value that minimized the SSE (lambda.min) rather than one standard error from lambda.min. It is because all coefficients under lambda.1se approach zero, shifting the model away from meaningful interpretation. Subsequent fixation of the regression model under lambda.min revealed the near-zero coefficients of quadratic terms associated with BMI and cortisol, indicating the insignificance of these quadratic variables in influencing DC values. Concurrently, LASSO regression introduced an L1 penalty term of coefficients to the SSE, and SSE minimization was performed. As shown in Figure [4], LASSO shrank coefficients of all quadratic terms to zero at lambda.min. This reiterated the conclusions drawn from Ridge Regression, reinforcing the inclusion of linear terms of variables and the interaction term between sex and cortisol. The successful alignment of results from penalized methods with manual variable selection underscores the robustness and consistency of our variable selection approach.

**Figure [3]:**
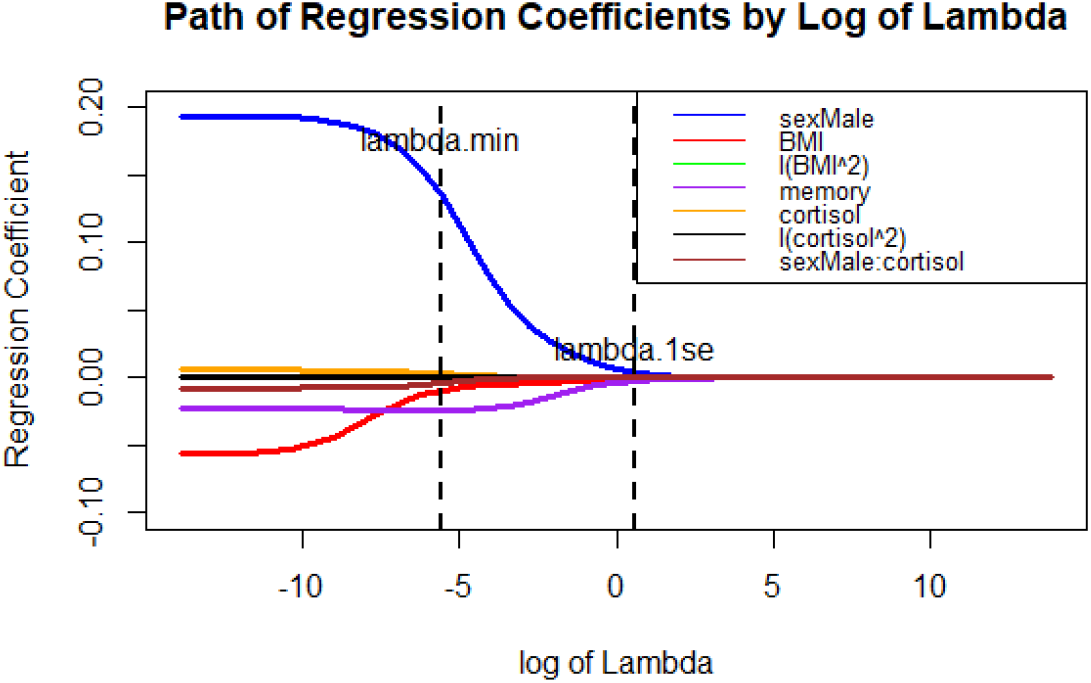
Ridge regression coefficients change by log of lambda.

**Figure [4]:**
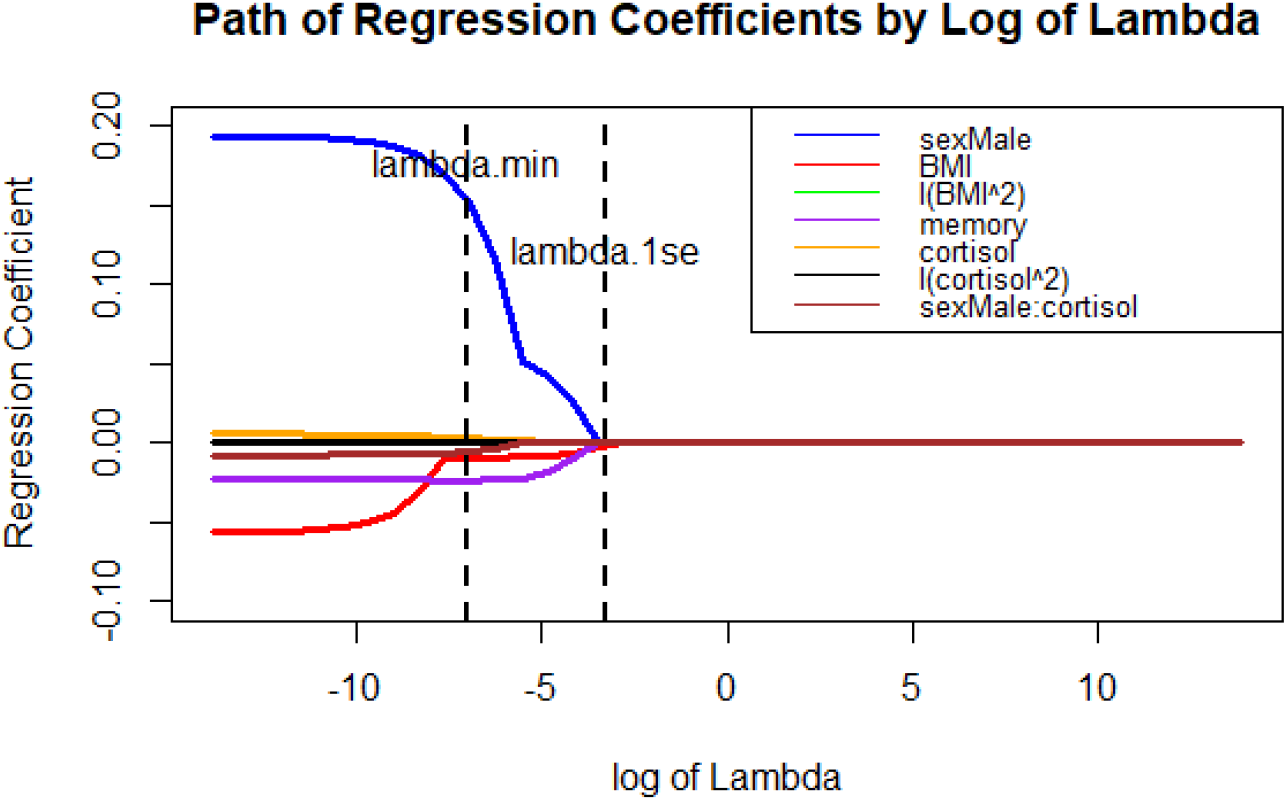
LASSO regression coefficients change by the log of lambda.

**Figure [5]:**
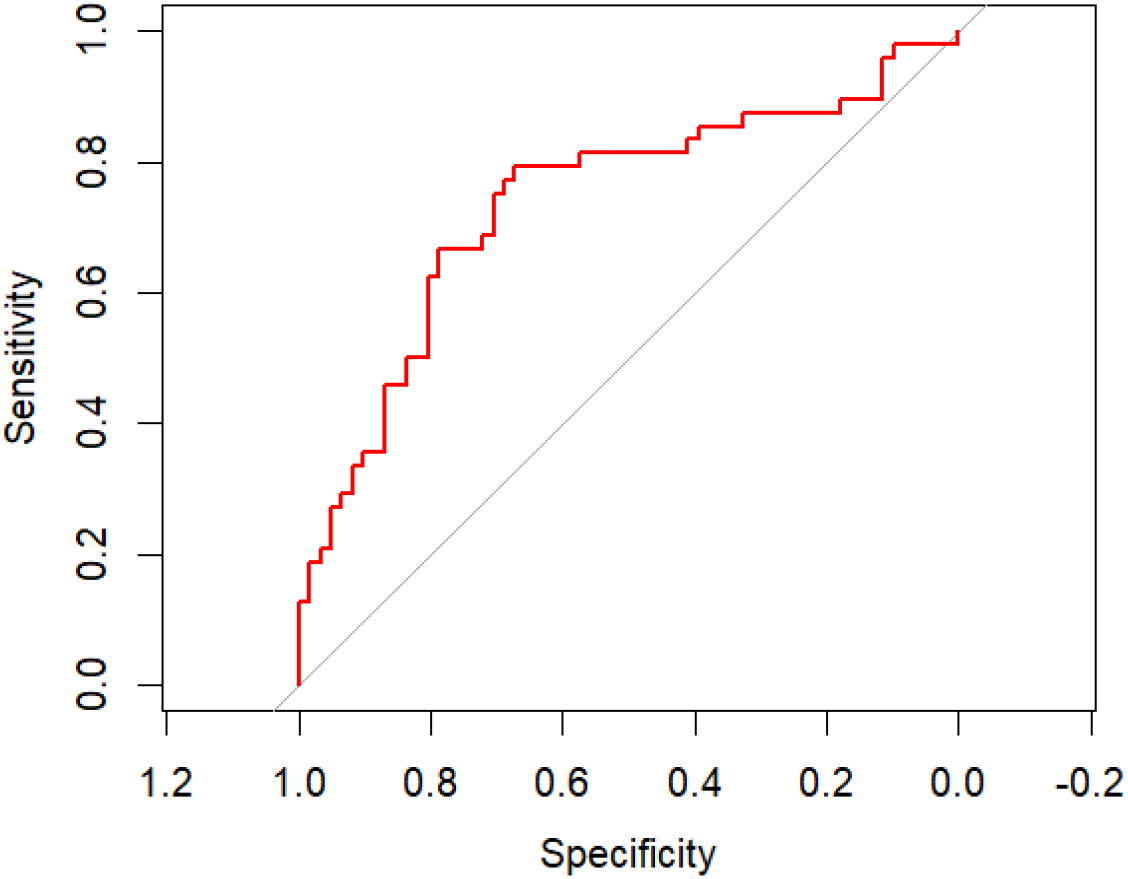
ROC curve plot of the final logistic model.

### Logistic Regression

Following the selected covariates, we constructed 3 nested models and evaluated their performance by AIC. One uses simple linear covariates for cortisol, sex, BMI, and numeric memory scores, one uses linear covariates with additional quadratic cortisol and BMI terms, and one uses linear covariates with additional quadratic cortisol and BMI terms, with memory score treated as factors. Again, the simplest logistic regression model with only linear covariate terms possessed the smallest AIC. In general, BMI and memory score were still negatively associated with the log odds ratio of high DC values vs. low DC values. Cortisol was positively associated with the log odds ratio of high DC values vs. low DC values.

Then, we proceeded to the model diagnostic process and obtained the influence plot of the final model. We found two additional observations that appear to be outliers. Observation 74 has a relatively low DC value, and Observation 16 has issues with both low DC value and extreme memory score.

In addition, we performed the goodness of fit test of our logistic regression model using the Hosmer-Lemeshow test.^11^ Since the p-value = 0.15, we failed to reject the null hypothesis and conclude that our model is adequately well-fitted. We also used the Receiver-Operator-Characteristics (ROC) curve to calculate the C-statistic, the probability a randomly selected patient with high DC values had a higher risk score than a patient with low DC values. Based on our ROC curve, we found that the area under the curve is 0.737, which indicates fairly good discrimination of the model^12^.

## Discussion

The increasing prevalence of “shidu” parents - those who have lost their only child - highlights a critical area of mental health concern, particularly the psychological impact and the risk of Post-Traumatic Stress Disorder (PTSD). Our study provides valuable insights into the complex relationship between Hippocampus Degree Centrality (DC) values and various risk factors in this population, which could contribute to more effective clinical interventions and support systems for these parents. By identifying key contributors such as cortisol levels, gender, BMI, and memory capability, we offer a predictive model that could guide psychiatric assessments and interventions.

Our findings emphasize the importance of a personalized approach when addressing PTSD in “shidu” parents. Specifically, the gender-specific correlation between cortisol levels and DC values suggests that males and females may respond differently to trauma. Males with lower cortisol levels appear more vulnerable to PTSD, while females with higher cortisol levels may experience more severe symptoms. These insights could be used to tailor interventions based on individual risk profiles, enhancing the effectiveness of psychological care for this group.

Additionally, BMI and memory scores were found to play significant roles in shaping hippocampal function, further underscoring the need to consider a wide range of physiological and cognitive factors when assessing PTSD risk.

While our models show strong potential for clinical application, they also underscore the complexity of PTSD, where multiple interacting risk factors contribute to outcomes. The models we developed, particularly the linear and logistic regression models, offer a framework for understanding these dynamics. By predicting DC abnormalities, which are indicative of stress levels and neurocognitive function, our approach provides a promising tool for early detection and intervention in “shidu” parents at risk of PTSD. Furthermore, these findings may guide policymakers in recognizing the broader public health implications of the one-child policy, particularly regarding mental health support for affected families.

However, there are limitations to our study. The relatively small sample size and the presence of missing data in a small subset of participants may reduce the generalizability and statistical power of our results. Furthermore, as our study focused exclusively on Han Chinese adults, the findings may not fully apply to other ethnic groups or populations with different cultural or socioeconomic contexts. Expanding future research to include larger, more diverse datasets would provide a more comprehensive understanding of PTSD in this population. Additionally, while our analysis employed multiple imputation techniques to address missing data, further refinement of data handling methods would improve the robustness of our models.

Future studies could build upon our findings by incorporating more advanced computational techniques, larger sample sizes, and a broader range of ethnic groups. By enhancing the predictive accuracy of PTSD risk models and considering other variables, such as genetic predispositions or environmental stressors, researchers could develop more personalized and effective interventions for “shidu” parents.

## Data Availability

The datasets used and/or analyzed during the current study are available from the corresponding author upon reasonable request. The code is available via https://github.com/Rub1sc0/Shidu.

## Acknowledgment

Sincerely thank Dr. Brent Coull for his invaluable feedback and insightful suggestions, which significantly improved the methods and results sections of this work.

## Footnotes

1 Pletcher, Kenneth. “one-child policy”. *Encyclopedia Britannica*, 7 Nov. 2023, https://www.britannica.com/topic/one-child-policy.

2 Xia, Zhuoman et al. “Relationship between SLC6A2 gene polymorphisms and brain volume in Han Chinese adults who lost their sole child.” BMC psychiatry vol. 24,1 11. 2 Jan. 2024, doi:10.1186/s12888-023-05467-4.

3 Du, Jun et al. “Post-traumatic stress disorder: a psychiatric disorder requiring urgent attention.” Medical review (2021) vol. 2,3 219-243. 2 Aug. 2022, doi:10.1515/mr-2022-0012.

4 Mondelli, Valeria et al. “Higher cortisol levels are associated with smaller left hippocampal volume in first-episode psychosis.” Schizophrenia research vol. 119,1-3 (2010): 75-8. doi:10.1016/j.schres.2009.12.021.

5 Ge, Jiyuan et al. “Persistence of post-traumatic stress disorder in Chinese Shidu parents is associated with combined gray and white matter abnormalities.” Psychiatry research. Neuroimaging vol. 335 (2023): 111715. doi:10.1016/j.pscychresns.2023.111715.

6 Xia, Zhuoman et al. “Relationship between SLC6A2 gene polymorphisms and brain volume in Han Chinese adults who lost their sole child.” BMC psychiatry vol. 24,1 11. 2 Jan. 2024, doi:10.1186/s12888-023-05467-4.

7 Not all individuals who lost their only child would develop PTSD or have changes in their hippocampus. The underlying mechanisms of PTSD development have not been adequately studied and understood.

8 Murray, Jared S. “Multiple Imputation: A Review of Practical and Theoretical Findings.” Statistical Science, vol. 33, no. 2, 2018, pp. 142–59, https://doi.org/10.1214/18-sts644.

9 Yamashita, Toshie, et al. “A Stepwise AIC Method for Variable Selection in Linear Regression.” *Communications in Statistics - Theory and Methods*, vol. 36, no. 13, 2007, pp. 2395–403, https://doi.org/10.1080/03610920701215639.

10 Wong, Ka Yiu, and Sung Nok Chiu. “An Iterative Approach to Minimize the Mean Squared Error in Ridge Regression.” *Computational Statistics*, vol. 30, no. 2, 2015, pp. 625–39, https://doi.org/10.1007/s00180-015-0557-y.

11 Hosmer, D.W., Hosmer, T. and Lemeshow, S. (1980) A Goodness-of-Fit Tests for the Multiple Logistic Regression Model. Communications in Statistics, 10, 1043-1069. https://doi.org/10.1080/03610928008827941

12 Sachs, Michael C. “plotROC: A Tool for Plotting ROC Curves.” *Journal of Statistical Software*, vol. 79, no. Code Snippet 2, 2017, pp. 1–19, https://doi.org/10.18637/jss.v079.c02.

## References

1. Abercrombie, Heather C et al. “Cortisol’s effects on hippocampal activation in depressed patients are related to alterations in memory formation.” Journal of Psychiatric Research vol. 45,1 (2011): 15–23. doi:10.1016/j.jpsychires.2010.10.005.

2. Du, Jun et al. “Post-traumatic stress disorder: a psychiatric disorder requiring urgent attention.” Medical review (2021) vol. 2,3 219–243. 2 Aug. 2022, doi:10.1515/mr-2022-0012.

3. Ge, Jiyuan et al. “Persistence of post-traumatic stress disorder in Chinese Shidu parents is associated with combined gray and white matter abnormalities.” Psychiatry research. Neuroimaging vol. 335 (2023): 111715. doi:10.1016/j.pscychresns.2023.111715.

4. Hosmer, D.W., Hosmer, T. and Lemeshow, S. (1980) A Goodness-of-Fit Tests for the Multiple Logistic Regression Model. Communications in Statistics, 10, 1043–1069. 10.1080/03610928008827941.

5. McAuley, M.T., Kenny, R.A., Kirkwood, T.B. et al. A mathematical model of aging-related and cortisol induced hippocampal dysfunction. BMC Neurosci 10, 26 (2009). 10.1186/1471-2202-10-26.

6. Mondelli, Valeria et al. “Higher cortisol levels are associated with smaller left hippocampal volume in first-episode psychosis.” Schizophrenia research vol. 119,1–3 (2010): 75–8. doi:10.1016/j.schres.2009.12.021.

7. Murray, Jared S. “Multiple Imputation: A Review of Practical and Theoretical Findings.” Statistical Science, vol. 33, no. 2, 2018, pp. 142–59, 10.1214/18-sts644.

8. Pletcher, Kenneth. “one-child policy”. Encyclopedia Britannica, 7 Nov. 2023, https://www.britannica.com/topic/one-child-policy.

9. Sachs, Michael C. “plotROC: A Tool for Plotting ROC Curves.” Journal of Statistical Software, vol. 79, no. Code Snippet 2, 2017, pp. 1–19, 10.18637/jss.v079.c02.

10. Wong, Ka Yiu, and Sung Nok Chiu. “An Iterative Approach to Minimize the Mean Squared Error in Ridge Regression.” Computational Statistics, vol. 30, no. 2, 2015, pp. 625–39, 10.1007/s00180-015-0557-y.

11. Xia, Zhuoman et al. “Relationship between SLC6A2 gene polymorphisms and brain volume in Han Chinese adults who lost their sole child.” BMC psychiatry vol. 24,1 11. 2 Jan. 2024, doi:10.1186/s12888-023-05467-4.

12. Yamashita, Toshie, et al. “A Stepwise AIC Method for Variable Selection in Linear Regression.” Communications in Statistics - Theory and Methods, vol. 36, no. 13, 2007, pp. 2395–403, 10.1080/03610920701215639.

